# Distinct neurofunctional alterations during motivational and hedonic processing of natural and monetary rewards in depression – a neuroimaging meta-analysis

**DOI:** 10.1101/2022.12.07.22283197

**Authors:** Mercy Chepngetich Bore, Xiqin Liu, Xianyang Gan, Lan Wang, Ting Xu, Stefania Ferraro, Liyuan Li, Bo Zhou, Jie Zhang, Deniz Vatansever, Bharat Biswal, Benjamin Klugah-Brown, Benjamin Becker

## Abstract

Reward processing dysfunctions are considered a candidate mechanism underlying anhedonia and apathy in depression. Neuroimaging studies have documented that neurofunctional alterations in mesocorticolimbic circuits may neurally mediate these dysfunctions. However, common and distinct neurofunctional alterations during motivational and hedonic evaluation of monetary (extrinsic) and natural (intrinsic) rewards in depression have not been systematically examined. Here, we capitalized on pre-registered neuroimaging meta-analyses to (1) establish general reward-related neural alterations in depression, (2) determine common and distinct alterations during the receipt and anticipation of monetary versus natural rewards, and, (3) characterize the differences on the behavioral, network and molecular level. The pre-registered meta-analysis (https://osf.io/ay3r9) included 633 depressed patients and 644 healthy controls and revealed generally decreased subgenual anterior cingulate cortex and striatal reactivity towards rewards in depression. Subsequent comparative analysis indicated that monetary rewards led to decreased hedonic reactivity in the right ventral caudate while natural rewards led to decreased reactivity in the bilateral putamen. These regions exhibited distinguishable profiles on the behavioral, network and molecular level. Further analyses demonstrated that the right thalamus and left putamen showed decreased activation during the anticipation of monetary reward. The present results indicate that distinguishable neurofunctional alterations may neurally mediate reward-processing alterations in depression, in particular, with respect to monetary and natural rewards. Given that natural rewards prevail in everyday life, our findings suggest that reward-type specific interventions are warranted and challenge the generalizability of experimental tasks employing monetary incentives to capture reward dysregulations in everyday life.

## Introduction

Pleasure and rewards constitute vital attractors that not only shape our momentary decisions, but also motivate future behavior. Deficits in reward processing have been proposed as a candidate mechanism underlying anhedonia, which describes a consistently diminished pleasure and interest in almost all daily life activities (Halahakoon et al., 2020), (Research Domain Criteria framework: positive valence system) (Insel et al., 2010). Anhedonia is a cardinal symptom of Major Depressive Disorder (MDD) and it represents a transdiagnostic symptom of several debilitating neurological and mental health disorders (Husain & Roiser, 2018). With currently over 320 million people living with depression, MDD has become a leading cause of disability worldwide (Friedrich, 2017). While the established interventions can alleviate some of the symptoms of depression, anhedonia is not only inefficiently targeted by the available treatments, but it is also highly resistant (Becker, 2022; Kendler, 2016; Robbins, 2016; Treadway & Zald, 2011). Understanding the specific dysfunctions of behavioral and neural reward processing in depression may thus provide better treatment and management option (Oh, Lee, Patriquin, Oldham, & Salas, 2021).

Reward processing encompasses an entire array of subprocesses that can be defined based on different perspectives including the class of external reinforcers as well as the stage and function of the rewarding process. Several stimuli can serve as rewarding reinforcers in humans, including money, palatable food and positive social interactions. These rewards share behavioral and neural features, such as motivational properties and engagement of the dopaminergic mesocorticolimbic pathways (Sescousse, Caldú, Segura, & Dreher, 2013), including activation of the ventral striatum (Mas-Herrero, Maini, Sescousse, & Zatorre, 2021). Monetary rewards are the most commonly used experimental paradigms in basic and psychiatric human neuroscience, but in addition to shared neural pathways (Ait Oumeziane, Schryer-Praga, & Foti, 2017; Flores, Münte, & Doñamayor, 2015), differential neural responses and alterations in patients with depression have been reported for monetary and naturalistic stimuli such as food and social rewards (Ait Oumeziane, Jones, & Foti, 2019; Rademacher et al., 2010; Spreckelmeyer et al., 2009; Yang et al., 2021).

Most fMRI incentive delay tasks employ monetary and social rewards that are generally considered to be secondary and learned, and can be differentiated from food as a primary reward, monetary rewards differ from all other rewards in important aspect. In particular, (a) they possess instrumental or exchangeable value and hence, can be used to acquire other desired goals, and (b) the receipt of monetary rewards commonly has a stronger association with the performance or efforts of the participant at least in the prevailing experimental paradigms (Sescousse et al., 2013). In contrast, natural rewards (often) do not possess an instrumental value and often have a less direct association with performance or effort. Although both reward types are strong motivational drivers in humans, natural rewards prevail in daily life and the pursuit of these rewards could be increased via therapeutic interventions in depression. The prevailing focus on monetary rewards in experimental studies may limit the generalizability and translation of altered reward-related brain processes in mental disorders, see (Ait Oumeziane et al., 2019) or (Zimmermann et al., 2019). Within this context, we focused on the comparison of monetary versus natural reward (encompassing palatable food and social rewards) processing alterations in depression.

On the neural level, reward processing has been closely linked to activation in mesocorticolimbic systems, including the striatum and amygdala as well as medial prefrontal and anterior cingulate regions (Rizvi, Pizzagalli, Sproule, & Kennedy, 2016; Schultz, 2015). While some evidence suggests that the specific neural systems vary as a function of the reinforcers and the stage of the reward process (Martins et al., 2021; Oldham et al., 2018; Yang et al., 2021), the ventral striatum has been consistently engaged during anticipation and receipt of monetary rewards (Jauhar et al., 2021), as well as the receipt of different natural rewards. According to overarching frameworks (Fehr & Camerer, 2007; Izuma, 2015), these meta-analytic findings may support the notion of a “common neural currency” for reward. However, (Knutson & Bossaerts, 2007) suggests phylogenetically different classes of rewards.

Previous meta-analyses have provided convergent evidence for reward processing deficits in depression (Halahakoon et al., 2020), with functional neuroimaging studies suggesting altered neural activity during reward processing (Keren et al., 2018; Yang et al., 2022). Alterations have been observed along the mesocorticolimbic reward circuit, including prefrontal regions such as the orbitofrontal cortex (OFC), medial prefrontal cortex (mPFC) and the anterior cingulate cortex (ACC), as well as subcortical regions including the striatum, amygdala, and hippocampus (Geugies et al., 2019; Keren et al., 2018; Ng, Alloy, & Smith, 2019; Russo & Nestler, 2013). Neurofunctional alterations in the striatum and ACC during monetary reward processing have been consistently observed during different stages of depression, including subthreshold depression (Stringaris et al., 2015), major depressive episodes (Keren et al., 2018) or remitted depression (Dichter, Kozink, McClernon, & Smoski, 2012), and may thus represent a common marker for depression-associated reward deficits. However, findings with respect to depression-related alterations during natural reward processing have revealed less consistent results (Oh et al., 2021; Smoski, Rittenberg, & Dichter, 2011). Several electroencephalography (EEG) studies that directly compared processing of monetary and natural rewards in depression suggest not only common but also domain-specific neurofunctional alterations (Ait Oumeziane et al., 2019; Nelson & Jarcho, 2021). Against this background, we examined if depression is characterized by a dysregulation of the ‘common neural currency’ for rewards by capitalizing on a robust fMRI meta-analytic strategy that allowed us to determine common and separable neurofunctional alterations during natural versus monetary reward processing and during anticipation versus receipt of reward in depression.

We conducted a preregistered coordinate-based meta-analysis according to the latest guidelines (Müller et al., 2018; Page et al., 2021) and included case-control fMRI studies examining the anticipation and receipt of monetary and natural rewards in depression. To facilitate a robust determination, we employed Seed-based d Mapping with Permutation of Subject Images (SDM-PSI), a novel method of performing neuroimaging meta-analyses (Albajes-Eizagirre, Solanes, Fullana, et al., 2019). SDM-PSI allows (nearly) unbiased estimation of effect sizes and generation of neurofunctional maps based on both positive and negative differences and hence can produce signed differential effect size maps. In line with the goals of the present study, we employed a three-step approach to meta-analytically determine common and domain-specific reward dysfunctions in depression, including a pooled analysis determining non-specific reward alterations, as well as separate meta-analyses determining common and domain-specific hedonic dysregulations during the receipt of monetary versus natural rewards or the anticipation and receipt of monetary rewards, respectively. We hypothesized domain-general alterations in the ventral striatal reward processing system (reflecting dysregulations in the ‘common neural currency’), but domain-specific alterations in prefrontal systems involved in more complex value and evaluation processes. To further characterize common and separable alterations, we employed a series of connectivity and meta-analytic strategies to distinguish the identified regions on the behavioral, network, and genetic levels.

## Method

### Data collection procedures, inclusion criteria and data extraction

The current pre-registered meta-analysis adhered to the guidelines of conducting coordinate-based neuroimaging meta-analyses (Müller et al., 2018). Procedures, hypotheses and analyses were pre-registered on the OSF-repository (https://osf.io/ay3r9). A comprehensive literature search was performed based on the Preferred Reporting Items for Systematic Reviews and Meta-Analyses (PRISMA) guidelines (Page et al., 2021). The search aimed to identify case-control fMRI studies examining reward processing in patients with depression in comparison to healthy controls. For the literature search, biomedical and life science databases including PubMed (https://www.ncbi.nlm.nih.gov/pubmed), Web of Science (https://www.webofscience.com/wos/alldb/basic-search) and PsycInfo (https://www.apa.org/pubs/databases/psycinfo/) were utilized.

Original case-control fMRI studies examining monetary and natural reward processing in depression were extracted and suitable studies from the reference lists of review articles were additionally included. The literature was independently screened based on titles and abstracts by M.C.B and B.K.B. The search terms were: “functional magnetic resonance imaging OR fMRI” AND “depression OR major depressive disorder OR unipolar depression OR sub-clinical depression OR at-risk of depression” in combination with either “monetary reward” OR “monetary incentive delay” OR “natural reward” OR “social reward”. Studies written in English, published from January 2001 to December 2021 and reporting whole brain results in standard stereotactic space (Talairach or Montreal Neurological Institute) were included. Only peer-reviewed, original case-control studies comparing patients and healthy controls were included.

Additional exclusion criteria were: (a) studies focusing entirely on participants <18 years and >60 years of age, (b) studies focusing on other mood disorders, including bipolar disorder and postpartum depression, (c) studies reporting region-of-interest (ROI) results only, (d) studies focusing on other reward-related processes such as probabilistic reward learning, prediction error etc., (e) studies reporting findings from an identical dataset as already included studies. The systematic literature review identified 26 suitable neuroimaging studies according to the inclusion criteria, 17 examined monetary reward and 10 examined natural reward (e.g., viewing positive images, positive social feedback, or chocolate) processing in depression.

### Aims and coordinate-based meta-analytic implementation

The present case-control neuroimaging meta-analysis aimed to segregate neurofunctional reward alterations in depression, in particular to disentangle general from domain-specific dysregulations in the domains of (a) monetary versus natural reward processing as well as in the domains of (b) anticipation versus receipt of rewards. In an initial step, general reward processing alterations were examined in an overarching meta-analysis that pooled the data from all studies (analysis 1). Next, our primary meta-analysis determined domain-specific alterations in hedonic processing by means of two separate meta-analyses comparing alterations during the receipt of monetary rewards versus natural rewards in depression (analysis 2). Finally, stage-specific alterations with respect to anticipation versus receipt processing of monetary rewards in depression were disentangled by another meta-analysis (analysis 3). However, the number of studies for anticipation of natural rewards was insufficient to explore domain-specific alterations between natural and monetary reward anticipation. All analyses were implemented in Seed-based d Mapping with Permutation of Subject Images (SDM-PSI) version 6.21 (https://www.sdmproject.com/) a novel and highly robust technique for performing neuroimaging meta-analyses (Albajes-Eizagirre, Solanes, Fullana, et al., 2019) (**supplements**).

To ensure independence of all studies in our meta-analysis, the coordinates of each study were put in a separate text file according to the guidelines of conducting meta-analysis using SDM-PSI. All analyses were thresholded at p ≤ 0.0025 uncorrected, k ≥ 10 voxels. This threshold was in line with the current meta-analysis and was chosen because it balances Type I and Type II errors (Chavanne & Robinson, 2021; X. Liu et al., 2022). Comparative analyses between monetary and natural reward outcomes were performed using Statistical Parametric Mapping (SPM12) software package (https://www.fil.ion.ucl.ac.uk/spm/). The analysis aims to find any common and separate areas of activation between the two identified clusters for natural and monetary reward alterations. We subjected the corresponding two statistical maps of NifTI images (of meta-analytic results of monetary reward outcome and natural reward outcome) to a direct comparison by means of the ImCalc function as implemented in SPM12 (**supplements**).

### Characterization of domain-specific hedonic dysregulations on the network, behavioral, genetic and receptor level

Given that our primary meta-analysis revealed distinct neurofunctional alterations during processing of monetary and natural rewards, we further characterized the identified signatures on the functional and neurobiological levels. Network analyses were performed using the Neurosynth database (Yarkoni, Poldrack, Nichols, Van Essen, & Wager, 2011) to determine whether the identified regions represent nodes of separable networks. This connectivity analyses were performed using peak coordinates of the caudate (MNI 10/10/4) and putamen (MNI22/8/4). Additional seed-to-whole-brain functional connectivity analyses from an independent dataset were conducted to provide a more fine-grained mapping of the common and separable intrinsic network organization of the identified striatal subregions (Zhao et al., 2019). Second, distinct behavioral functions of the regions were identified through meta-analytic topic mapping using peak coordinates of the identified brain regions from the Brain annotation toolbox (BAT) (Z. Liu et al., 2019). Third, to examine separable genetic underpinnings of the identified signatures, we examined genetic expressions of these regions using the Brain annotation toolbox (BAT). This determined which genes had the highest densities and separable expressions in the identified regions (see **supplements**).

### Sensitivity, Heterogeneity and Publication bias

Inter-study heterogeneity for each cluster was examined by the *I^2^* index which represents the proportion of the total variation caused by study heterogeneity (Higgins & Thompson, 2002). Generally, *I^2^*> 50% indicates substantial heterogeneity (Martins et al., 2021). Jackknife sensitivity analysis (repeating the meta-analysis after discarding one study each time) assessing the robustness of the meta-analysis was conducted on Anisotropic Effect Size-Signed Differential Mapping (AES-SDM). Funnel plots (asymmetrical funnel plots indicate that results were driven by few studies or studies with a small sample size, while symmetric plots suggest otherwise) and Egger’s tests checked for publication bias (an asymmetric plot and p<0.05 were recognized as significant).

### Exploratory analyses of potential confounders - Linear Model analysis

A linear model analysis to determine the potential effects of medication on neurofunctional alterations of monetary and natural outcome. Medication and non-medication studies were coded as 1 and 0, respectively. Moreover, we repeated the meta-analysis including only unmedicated studies to confirm the stability of the results, i.e., whether the results remain stable without medication. Previous studies have shown that age and gender affect reward-related brain activity (Greimel et al., 2018). Meta-regressions were performed to explore the influences of other potential confounders.

## Results

The comprehensive literature search resulted in 26 suitable original fMRI studies on reward processing in depression, i.e., 17 monetary reward studies and 10 natural reward studies (8-social reward and 2-food reward), with n=1277 participants (n=633 patients, n=644 healthy controls). The database included patient data from monetary (*n*=391, mean age=31.47, *SD*=8.41) and natural reward studies (*n*=242, mean age=35.04, *SD*=10.26). There were no significant differences in age (p=.21, t=2.07) and gender (p=.97, t=2.10). Healthy controls data from monetary reward studies (*n*=393, mean age=30.95, *SD*=7.89) and natural reward studies (*n*=251, mean age=31.9, *SD*=7.98) had no significant differences in age (p =.69, t=2.08) and gender (p=.53, t=2.26). **Fig. 1** shows the flow diagram of the selection process. Demographic information of the included studies is displayed in **Table 1** (**Supplementary Table S1** - characteristics of included studies and **Supplementary Table S2** - PRISMA checklist).

**Fig. 1.**
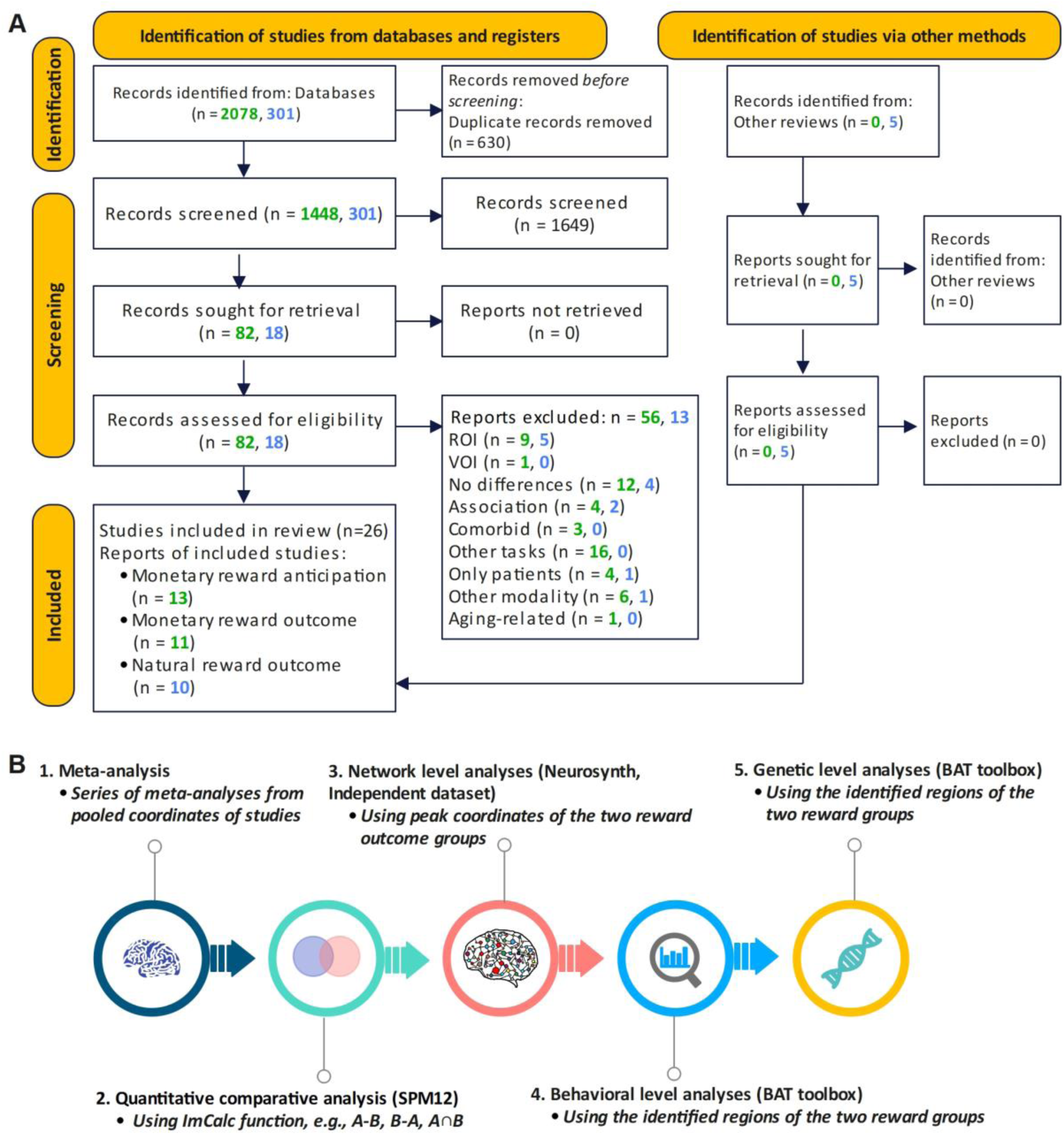
Flow diagram of the systematic literature search and identification of suitable original studies.

**Table 1.**
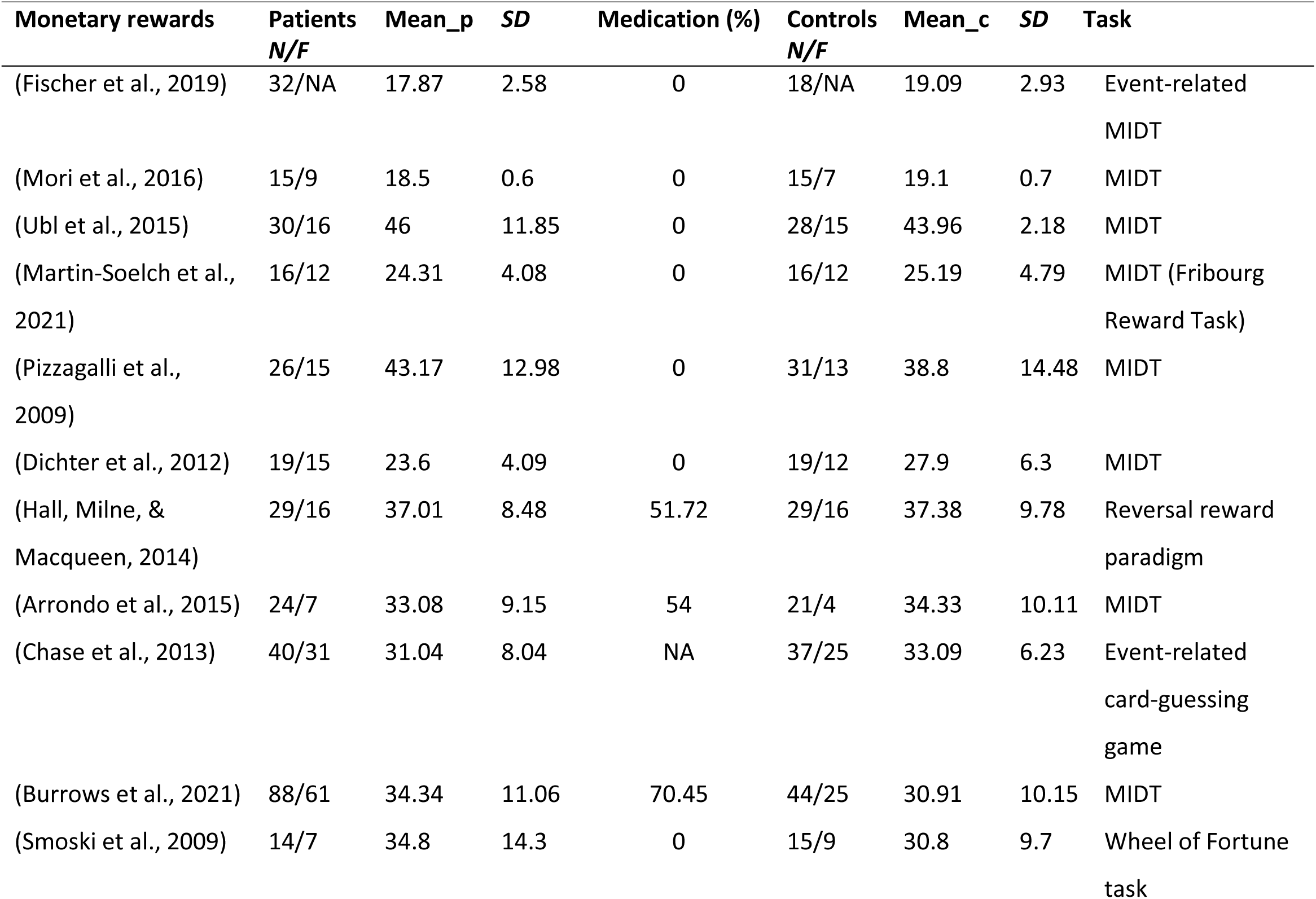
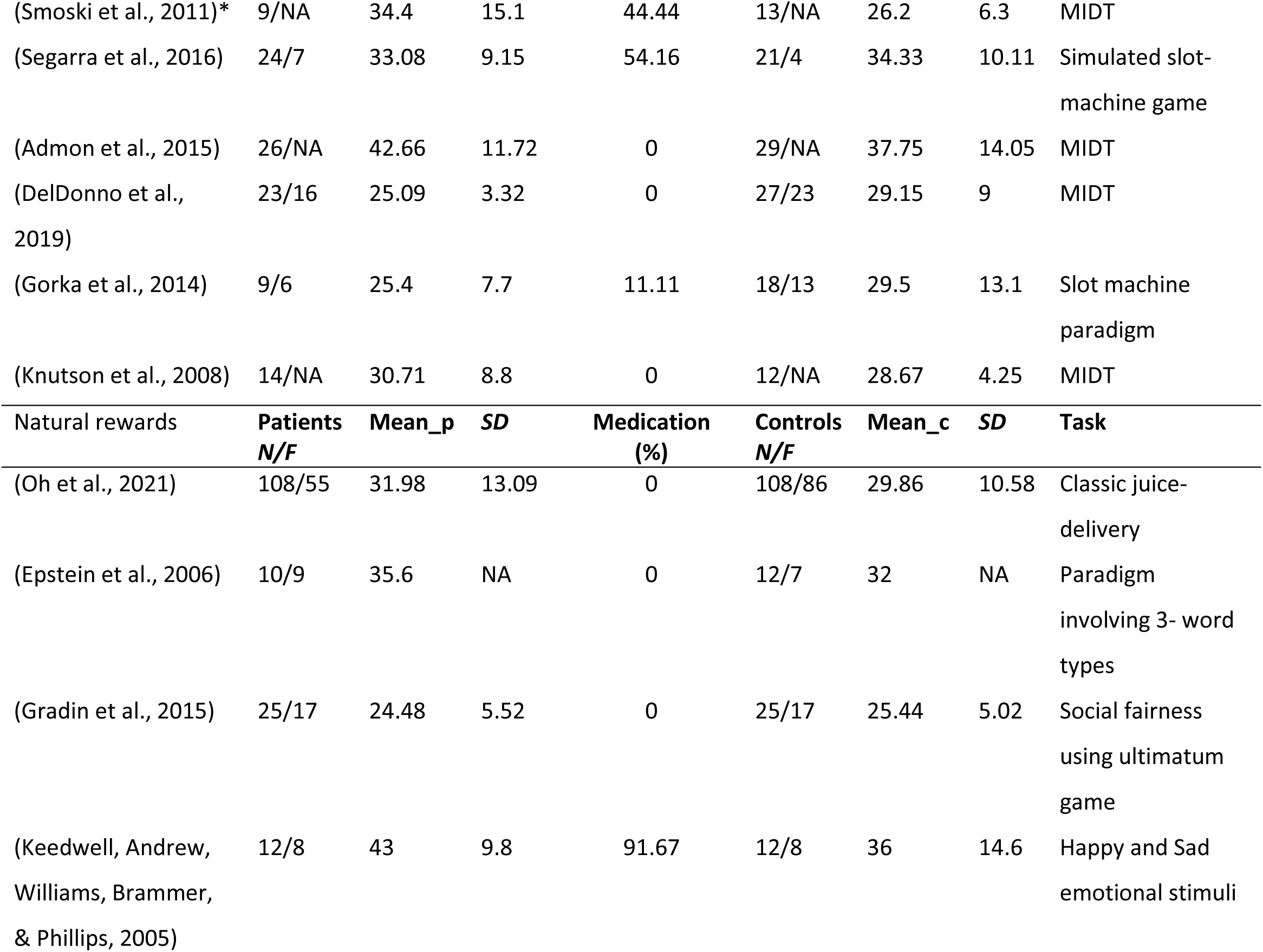
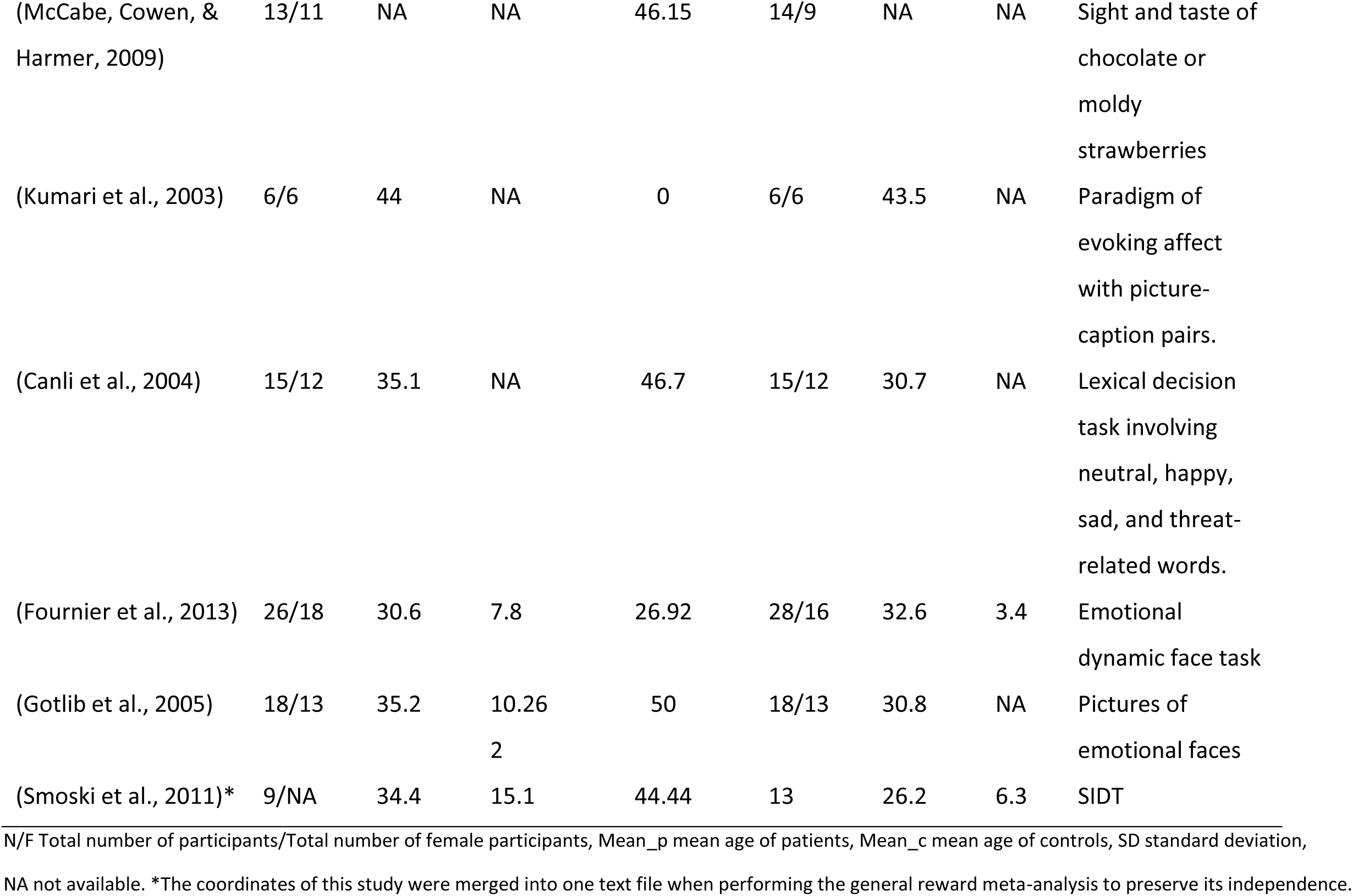
Demographic information of included studies.

### Neurofunctional reward alterations in depression

#### Analysis 1: General reward processing alterations in depression

Meta-analytic results obtained from the pooled studies of monetary and natural reward outcome revealed decreased activation in the right striatum, with the cluster encompassing mainly the putamen and extending into the caudate, and the subgenual anterior cingulate in depression (**Fig. 2**). Between-study heterogeneity and publication bias are displayed in **Table 2** (**Supplementary Figure S1**-funnel plots).

**Fig. 2.**
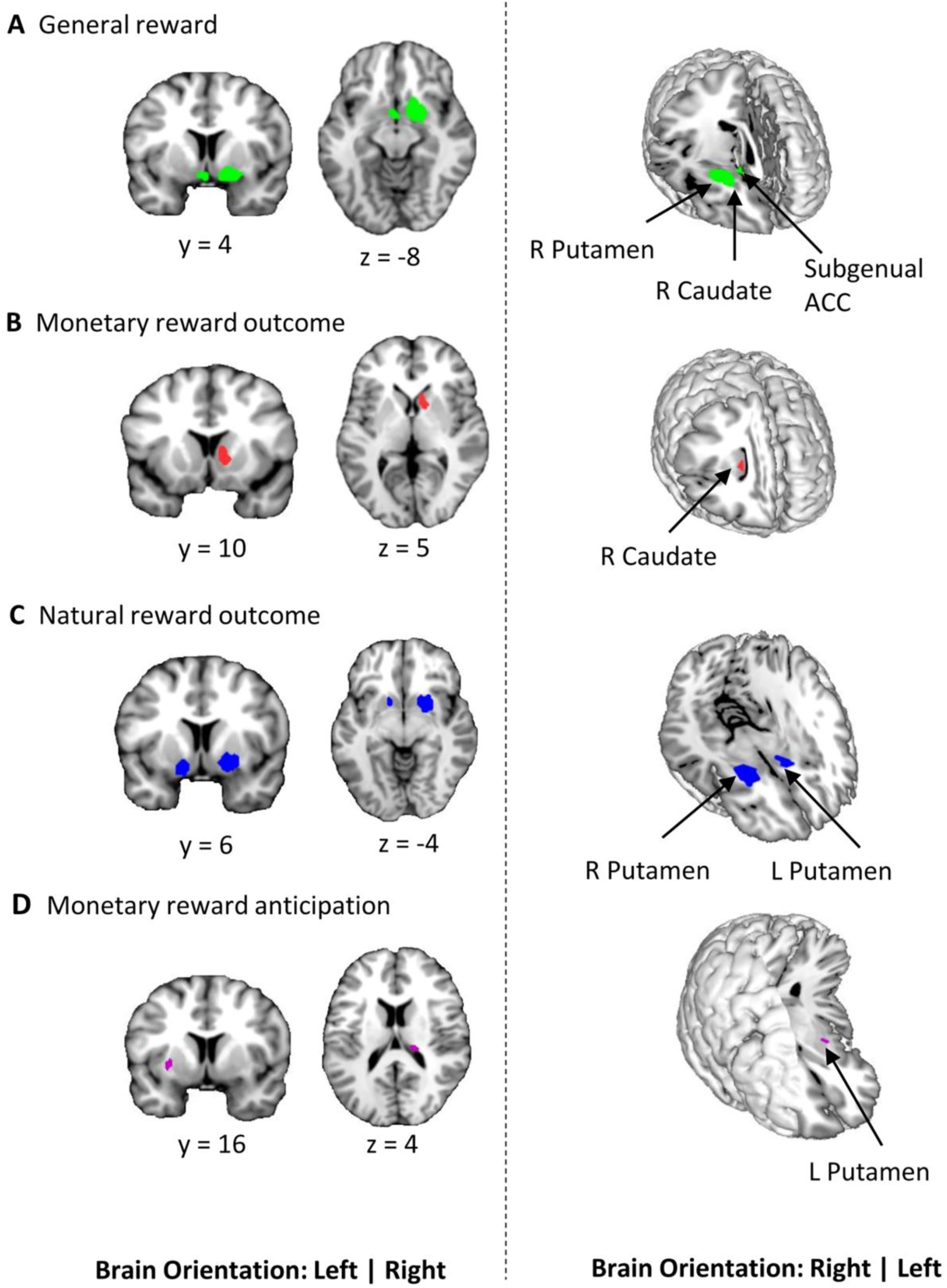
Results of the primary meta-analyses on dysfunctional reward processing in depression. **A** Display of regions that showed generally reduced reactivity during reward processing in depression as compared to healthy controls. Subsequent meta-analyses revealed regions that are specifically dysfunctional during the hedonic processing of **B** monetary and **C** natural rewards. **D** shows region with altered activation in depression during the anticipation of monetary rewards (p < 0.0025 uncorrected, k ≥ 10 voxels).

**Table 2.** Whole-brain meta-analysis results fMRI studies in unipolar depression at *p* < 0.0025 uncorrected threshold.

| Reward type | MNI peak coordinates | Main cluster | SDM Z | Voxels | Hedges' g | $I^2$ statistic | Egger's $p$-value | Jackknife sensitivity |
| --- | --- | --- | --- | --- | --- | --- | --- | --- |
| General rewards | Depression < HC |  |  |  |  |  |  |  |
|  | 14, 4, -8 | R caudate/putamen | 4.615 | 294 | -0.42 | 6.76% | 0.55 | 20/19 |
|  | 0, 2, -8 | Subgenual ACC | 3.740 | 35 |  | 2.55% | 0.85 |  |
| Monetary outcome | Depression < HC |  |  |  |  |  |  |  |
|  | 10, 10, 4 | R caudate | 4.260 | 87 | -0.58 | 11.75% | 0.57 | 11/10 |
| Natural outcome | Depression < HC |  |  |  |  |  |  |  |
|  | 22, 8, -4 | R putamen | 4.826 | 223 | -0.57 | 2.08% | 0.95 | 11/10 |
|  | -14, 8, -6 | L putamen | 4.164 | 143 | -0.52 | 0.70% | 0.89 |  |
| Monetary anticipation | Depression < HC |  |  |  |  |  |  |  |
|  | 18, -26, 12 | R thalamus | 3.214 | 18 | -0.39 | 18.63% | 0.61 | 14/13 |
|  | -30, 4, -2 | L putamen | 3.466 | 12 | -0.37 | 4.17% | 0.99 |  |

ACC anterior cingulate cortex, fMRI functional magnetic resonance imaging, HC healthy controls, MNI Montreal Neurological Institute, L left hemisphere, R right hemisphere, SDM seed-based d mapping.

#### Analysis 2: Domain-specific (monetary versus natural) reward processing alterations in depression

During monetary reward outcome, depressed patients exhibited decreased activation in the right caudate compared to healthy controls (**Fig. 2B**). Depressed patients exhibited decreased activation in more dorsal parts of the striatum with the clusters being specifically located in the bilateral putamen during natural reward outcome (**Fig. 2C**). For a more fine-grained localization of the monetary versus natural reward processing alterations, additional mapping using the Human Brainnetome Atlas (Fan et al., 2016) was employed. Peak coordinates of the monetary outcome alterations (MNI 10/10/4) mapped onto the ventral caudate whereas coordinates for the natural reward alterations (MNI 22/8/-4) mapped onto the ventromedial putamen supporting a ventral to dorsal striatum dissociation of the alterations (Zhao et al., 2019; X. Zhou et al., 2019).

#### Analysis 3: Stage-specific (anticipation versus outcome) reward processing alterations in depression

There was decreased activation in the right thalamus and left putamen during anticipation (**Fig. 2D**) in depression relative to controls. Patients displayed decreased activation in the right caudate during monetary reward outcome. There were no regions of increased activation across all four meta-analyses.

### Domain-specific (natural vs monetary) reward dysregulations: regional comparative, network level and behavioral characterization

According to the aims of the present study, comparative analyses were implemented to further determine common versus separable regions of altered monetary vs natural reward outcome processing in depression. The direct comparison did not reveal an overlap between neural alterations in the two domains. It confirmed that decreased monetary outcome activity was located in the right caudate. In contrast, decreased natural outcome alterations were located in more dorsal regions of the striatum, i.e., the bilateral putamen (**Fig. 3A**).

**Fig. 3.**
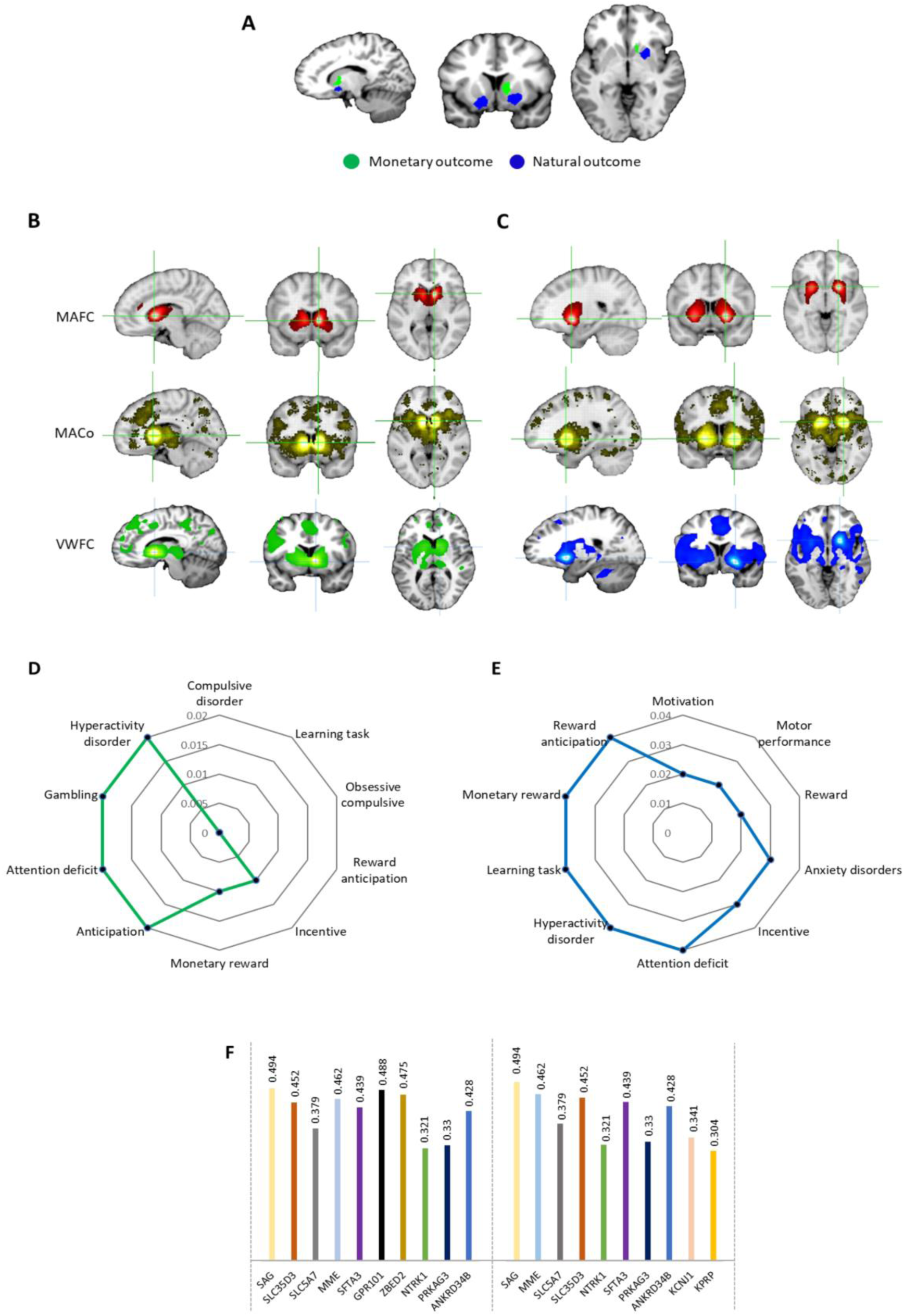
Regional, behavioral, network, and molecular level characterization of the monetary and natural reward outcome alterations in depression. **A** Comparative analysis of monetary versus natural outcome alterations in depression. Further meta-analytic and voxel-wise characterization of the identified regions using Neurosynth and independent resting state fMRI data. **B** Brain networks of the identified caudate region of monetary reward outcome alterations. **C** Brain networks of the identified putamen region of natural reward outcome alterations. MAFC, meta-analytic functional connectivity; MACo, meta-analytic co-activation; VWFC, voxel-wise functional connectivity. **D** Meta-analytic topic mapping of the right caudate. **E** Meta-analytic topic mapping of the right putamen. The terms are arranged and read from top in clockwise direction according to p-values. **F** Genetic expression of the caudate (left side) and putamen (right side). The plot shows the genetic symbols according to their respective correlation values.

Network-level and co-activation characterization of the identified striatal regions for monetary (**Fig. 3B**) and natural reward dysregulations (**Fig. 3C**) revealed functional connectivity, meta-analytic co-activation, and voxel-wise functional connectivity maps. The caudate was identified for monetary reward outcome dysfunctions coupled with core regions of the mesocorticolimbic pathways, including bilateral ventral striatal regions, ventral tegmental area (VTA) and cortical midline structures, while the putamen was identified for natural reward outcome dysfunctions coupled stronger with bilateral dorsal striatal regions as well as lateral frontal and insular regions (**Supplementary Figure 2**).

Examination of the top 10 behavioral terms from the behavioral analyses revealed that both regions were characterized by reward, motivation and emotional arousal associated terms, while the caudate region was strongly related to decision making and compulsive behavior and the putamen region was stronger involved in motor and behavioral control (**Fig. 3D** and **Fig. 3E**, stronger associations are depicted as smaller p-values).

### Genetic level characterization

Genetic expression analyses revealed that S-antigen visual arrestin (SAG) was the most expressed gene in both caudate and putamen. The top ten genes were generally highly similar (**Fig. 3F**).

### Heterogeneity, publication bias and sensitivity

The results of tests for heterogeneity (*I^2^*), approximate effect sizes (Hedge’s *g*), Egger’s probabilities of publication bias as well as jackknife sensitivity results are presented in **Table 2**.

There was low heterogeneity across the main meta-analyses. Publication bias failed to reach statistical significance.

### Exploratory analyses

Additional control analyses revealed no consistent or strong evidence for the influence of potential confounders i.e., medication, age and gender (**supplements**).

## Discussion

Depressive mood and anhedonia represent key symptoms of major depression and have been associated with neurobiological dysregulations in reward and motivational processes. Many studies have utilized case-control fMRI designs to map the neurofunctional basis of reward dysregulations in individuals with depression. Several of these studies revealed altered neural activity in the mesocorticolimbic reward circuits in depression. However, it currently remains unclear whether the neurofunctional alterations vary as a function of the reward domain, i.e., natural rewards versus monetary rewards. We hereby conducted a pre-registered neuroimaging meta-analysis to systematically determine common and separable reward dysfunctions in depression. Quantitative analyses of 26 suitable neuroimaging studies focusing on reward-related processes in 633 patients and 644 controls revealed that the right striatum and subgenual ACC exhibit generally reduced reward reactivity in patients with depression, with subsequent comparative analysis demonstrating that more ventral parts of the right striatum (caudate) show reduced reactivity during monetary reward outcome while more dorsal and bilateral parts of the striatum (putamen) show reduced reactivity during the receipt of natural rewards. Additional meta-analytic characterization and an independent fMRI dataset revealed that the two identified regions exhibit common as well as separable characteristics. On the network level, the caudate interacted with ventral striatum, ventral tegmental area (VTA), and cortical midline structures while the putamen exhibited stronger connectivity with more dorsal striatal and lateral fronto-insular regions. On the behavioral and molecular level, both regions were characterized by similar genetics and an involvement in reward-related processes. However, the putamen showed a stronger involvement in cognitive and motor control related processes. Finally, an exploratory meta-analytic examination of stage-specific reward alterations yielded decreased activation in the right thalamus and left putamen during monetary reward anticipation in depression.

### Common alterations - general hedonic reward processing alterations in depression

The current meta-analysis revealed that the subgenual ACC and right putamen extending to the caudate, exhibited decreased reactivity in depression during general processing of reward. Our results align with several original studies that have demonstrated altered brain activity in these regions during reward processing in depression. The striatum has been associated with both, monetary and natural reward processing in healthy individuals (Pizzagalli et al., 2009) and dysfunctional reward processing in this region has been reported in several mental disorders, including depression, schizophrenia and addiction (Klugah-Brown et al., 2020; Pizzagalli et al., 2009; Zimmermann et al., 2019).

The subgenual ACC has been consistently involved in reward processing (Rogers et al., 2004), as well as regulatory control in cognitive and emotional domains (Critchley, 2005; Drevets, Savitz, & Trimble, 2008), and may serve an integrative function to evaluate rewards and guide the most appropriate choices between rewards (Holroyd & Yeung, 2012). Recent neuropathological models suggest a key role of the subgenual ACC in mood disorders including depression (Drevets, Price, & Furey, 2008). It is described as an interface system between the cognitive and emotional networks of the brain that conveys emotional information (Greicius et al., 2007). A previous meta-analysis has also shown decreased volume of the subgenual ACC in mood disorders (Hajek, Kozeny, Kopecek, Alda, & Höschl, 2008). Reduced activation in this region is distinctively associated with depression as it cuts across behavioral, emotional and cognitive aspects.

### Distinct neurofunctional alterations during hedonic processing of natural and monetary rewards

While our findings on the neurofunctional basis of general reward processing alterations in depression align with previous meta-analyses, the key aim of the present meta-analysis was to further segregate neurobiological dysregulations during hedonic processing of different rewards. We hypothesized a common ventral striatal deficit across natural and monetary rewards in depression, however we observed non-overlapping alterations. The ventral caudal regions displayed reduced reactivity during monetary rewards while more dorsal striatal regions showed decreased reactivity towards natural rewards. Both regions have been involved in reward processing across domains (Arsalidou, Vijayarajah, & Sharaev, 2020) and general reward processing alterations in depression (Keren et al., 2018). However, the striatum is organized along a ventral to dorsal axis with respect to different behavioral domains and network-level interactions. The ventral parts of the caudate have been primarily involved in reward, reinforcement and incentive salience processing while the more dorsal parts of the striatum mediate sensorimotor processes and some executive functions such as inhibitory control (Haber, 2016; Robbins, Gillan, Smith, de Wit, & Ersche, 2012; Zhuang et al., 2021). In line with this functional specialization of the striatum subregions, the identified caudate region exhibited strong connections with ventral parts of the striatum, the VTA and cortical midline structures strongly involved in reward-related processing while the putamen region coupled with dorsal striatal regions as well as lateral frontal and insular regions involved in interoceptive processes, pain empathy and regulatory control (Craig, 2009; F. Zhou et al., 2018; Zhuang et al., 2021). The network level differentiation was additionally mirrored on the behavioral level. Both regions contributed to reward processes, but the more ventral region was also involved in compulsivity, learning and motivational processes while the dorsal region showed additional involvement in motor and behavioral control processes. Together, the neurofunctional, network and behavioral characterization may indicate that, (1) different neurobiological mechanisms underlie depression-related reward dysregulations in the hedonic processing of monetary and natural rewards, and (2) a limited generalization and ecological validity of the prevailing reward paradigms incorporating small amounts of monetary incentives to situations in everyday life that engage primarily natural rewards.

On the molecular level, both regions exhibited a similar genetic profile with S-antigen visual arrestin (SAG), SLC35D3 and MME showing high expressions. SAG has been associated with the G-protein-coupled receptor reactivity and thus sensitivity to several neurotransmitters. SLC35D3 gene has been associated with mid-brain and striatal dopamine signaling (Zhang et al., 2014) while MME drives the inactivation of neuropeptides like oxytocin (Sterchi, Naim, Lentze, Hauri, & Fransen, 1988). Dopamine and oxytocin have been associated with network-level modulation of striatal circuits across reward domains (Grimm et al., 2020; Scheele et al., 2013; Zhao et al., 2019) and this may shape context-dependent reward dysregulations in depression.

### Stage-specific neurofunctional reward processing alterations in depression – anticipation vs outcome

While the outcome of rewards is commonly considered a hedonic process, anticipation of rewarding outcomes is mostly considered a motivational process. Dysregulated motivational and approach tendencies towards pleasant events have long been associated with depression (Davidson, 1998). However, previous findings in depression have remained largely inconsistent with respect to the anticipation of rewards. For instance, depressed patients have demonstrated elevated activation during monetary reward anticipation in the anterior cingulate (Dichter et al., 2009; Knutson, Bhanji, Cooney, Atlas, & Gotlib, 2008) while other studies reported a relatively decreased anticipatory signal in striatal regions including the bilateral caudate, bilateral putamen, ventral striatum and globus pallidus (Keren et al., 2018; Stringaris et al., 2015; Takamura et al., 2017). Partly resembling these previous findings, we observed decreased anticipatory activity during monetary rewards in the thalamus and putamen of depressed patients. The thalamus represents an early detection node for salient stimuli (Cho et al., 2013). Decreased striatal activity during anticipation of reward has been reported in depression (Pizzagalli et al., 2009), plus other mental disorders (Leroy et al., 2020; Luijten, Schellekens, Kühn, Machielse, & Sescousse, 2017) and thus may represent a transdiagnostic marker for deficient motivation.

The present findings have some limitations. First, the exploratory linear model analysis revealed some evidence for potential confounding effects of treatment on the identified region during monetary outcome (Delaveau et al., 2011). Although both reward results remained stable after excluding studies with medication, future studies are required to explore effects of treatment on reward dysregulations in depression. Second, some studies mixed unmedicated and medicated patient groups in a single study and this increases the chances of type II errors (Dichter et al., 2009). Third, the number of studies for natural reward anticipation was insufficient. Fourth, the number of studies was moderate, only 10 studies were included in the natural outcome category. Fifth, this meta-analysis focused on comparing monetary and natural rewards unlike previous studies which differentiated primary and secondary rewards (Sescousse et al., 2013), and another suggested distinguishable electrophysiological neural signatures between monetary, social and food rewards (Banica, Schell, Racine, & Weinberg, 2022).

However, the limited number of case-control fMRI studies on natural reward processing in depression did not allow us to further segregate primary versus secondary or specific secondary reward categories, respectively. It is difficult to determine whether the neurofunctional alterations determined during natural reward processing are held across all types of natural rewards included and also which natural reward processes deficits might be specifically relevant to depression. Moreover, the monetary and natural rewards may differ in other important aspects relevant for reward processing, including the magnitude of reward. Finally, some included articles did not explicitly present non-significant results and this may affect meta-analytic results (Albajes-Eizagirre, Solanes, & Radua, 2019). The present meta-analysis focused on the differential case-control contrasts (reward vs non-reward) provided by the original studies. For specificity of subsequent meta-analysis, it would be helpful if future studies also report results from within-group comparisons.

Summarizing, the present meta-analysis supports the notion of general reward processing deficits in depression. However, the underlying neurofunctional dysregulations vary according to reward type and sub-processes. Previous work has largely focused on monetary rewards to map disrupted reward processing in depression. However, this may have overshadowed deficits in the processing of natural rewards. Natural rewards prevail in everyday life and shape our social interactions and thus dysregulations in this domain may lead to marked impairments in everyday life. The present meta-analysis has revealed distinguishable neurofunctional reward processing deficits in depression, suggesting that natural and monetary reward processing alterations may be rooted in distinct neurobiological dysfunctions.

## Data availability

Studies included in the meta-analyses have been cited in the manuscript. Coordinates will be made available upon request.

