## Supplementary material for "Distinct neurofunctional alterations during motivational and hedonic processing of natural and monetary rewards in depression – a neuroimaging meta-analysis" (from another version of this preprint: version 3, 2023-07-26)

### Supplementary methods

The SDM-PSI approach followed the following steps: (a) Peak coordinates of between-group differences (patients versus healthy subjects) and their effect sizes in terms of either t/z values were extracted in accordance with the SDM inclusion criteria. Z-values representing coordinates from whole-brain between-group differences of depressed patients and healthy controls were transformed into t-values using the statistical converter (https://www.sdmproject.com/utilities/?show=Coordinates). (b) For studies reporting peak coordinates, the initial preprocessing step (Anisotropic full width half maximum (FWHM) = 20mm and voxel size = 2mm) estimated the lower and upper bounds (Hedges' g) of the most probable effect size images (Radua et al., 2014). (c) The mean is analyzed by the maximum-likelihood estimation (MLE) and meta-analysis of non-statistically significant unreported effects (MetaNSUE) algorithm. This not only estimates the most likely effect size and the standard error, but it also creates numerous imputations based on the estimates that are within the bounds (Albajes-Eizagirre, Solanes, & Radua, 2019). (d) Next, imputed study images are recreated. (e) Finally, the permutation test evaluated the combined meta-analysis images for statistical significance. The generated maps were visualized using multi-image analysis GUI (MANGO; http://ric.uthscsa.edu/mango). Furthermore, inter-study heterogeneity for each cluster was examined by the $I^2$ index which represents the proportion of the total variation caused by study heterogeneity (Higgins & Thompson, 2002). Generally, $I^2$ values have been categorized into three groups namely low (25%), moderate (50%) and high (70%) (Martins et al., 2021).

#### *Comparative analysis*

The statistical maps of monetary and natural reward outcome alterations in MDD versus healthy controls were subjected to the SPM12 ImCalc function. Three calculations were performed, i.e., A-B (Monetary outcome – Natural outcome), B-A (Natural outcome -

Monetary outcome) and A∩B (the overlap activated brain region of monetary and natural reward outcome).

*Analyses on the network, behavioral, genetic and receptor level*

First, meta-analytic network analyses were utilized to determine whether the identified regions represent nodes of separable networks. Functional decoding was conducted using the Neurosynth database (Yarkoni, Poldrack, Nichols, Van Essen, & Wager, 2011) (https://www.neurosynth.org/). Peak coordinates of the identified signatures were used to generate region-specific unthresholded resting-state functional connectivity maps, meta-analytic co-activation maps and their respective conjunction maps. The resultant resting-state functional connectivity maps were thresholded at r > 0.2. The thresholded map was transformed into z-scores. Next, conjunction maps between the thresholded resting state functional connectivity and meta-analytic co-activation maps were generated for each ROI signature using SPM and the resultant regions were identified with the Anatomy toolbox (Eickhoff et al., 2005). The functional connectivity maps represent brain regions co-activated across the resting-state fMRI time series with the seed regions. Meta-analytic co-activation maps symbolize co-activation of brain regions across all fMRI studies in the Neurosynth database. The combination of functional connectivity and meta-analytic co-activation maps allows for analyzing both task-independent and task-driven functional networks emerging from the seed region.

Additional seed to whole brain functional connectivity analyses on the voxel level were conducted to provide a more fine-grained mapping of the common and separable intrinsic network organization of the identified striatal subregions (Zhao et al., 2019). To this end, data from an original independent study was included. To conduct functional connectivity analyses, data was collected from the Open Access Series of Imaging Studies (OASIS)-3 dataset (https://central.xnat.org) including n = 100 healthy subjects. Preprocessing was performed using Data Processing Assistant for Resting-State fMRI (http://rfmri.org/DPARSF). Briefly, functional imaging preprocessing procedures consisted of the following steps: (a) Deletion of the initial 5 volumes of the time-series, (b) Motion correction using rigid body translation and rotation and exclusion of subjects with maximum motion > 2 mm or 2°, (c) Two-step procedure of normalization into standard stereotactic space including co-registration to anatomical images and application of the corresponding segmentation matrix to the functional time series. 6mm radius spheres centered at the corresponding peak coordinates (caudate: x=10, y=10, z=4 and putamen: x=22, y=8, z=-4) served as seed regions for the seed-to-whole-brain voxel-wise analyses implemented in Data Processing Assistant for Resting-State fMRI (http://rfmri.org/DPARSF).

Behavioral-level characterization was achieved by obtaining the top twenty behavioral terms together with their p-values values (permutation p values arranged in descending order, where $p_{perm} < 0.05$ is significant). For the genetic level characterization, the top ten genes were acquired in terms of their correlation values. Using PubMed gene (https://www.ncbi.nlm.nih.gov/gene), we determined the identified genes' main functions.

**Supplementary results**

*Exploratory analyses*

Meta-regression analyses revealed that the results obtained from the main meta-analysis were not influenced by age or gender (**Supplementary Figure S4**). Linear model analyses further revealed that medication had no significant effect on the alterations observed during natural reward outcome (left and right putamen). However, the linear model for medication effects during monetary outcome overlapped with the meta-analytic effects of depression identified in the right caudate (**Supplementary Figure S5**). To further determine a potential confounding effect of medication, we recomputed the analyses after excluding data from patients with medication and differences between patients and healthy controls in terms of decreased activity during monetary reward outcome remained stable in the right caudate (retaining data from 7/11 studies on monetary reward outcome, *p* < 0.0025).

**Table S1.** Characteristics of included studies

| Study | Space | Correction | Contrast | Treatment |
|---|---|---|---|---|
| (Fischer et al., 2019) | MNI | p < .05 | Controls>At-risk, Anticipation: Reward>Loss Outcome: Reward>Loss | Unspecified. |
| (Mori et al., 2016) | MNI | Uncorrected at p < .001 | Subthreshold depression>Controls, Anticipation: Gain Vs No gain | Unmedicated at time 1. |
| (Ubl et al., 2015) | MNI | Corrected at p <.05 | Controls>Patients, Anticipation | Unmedicated. |
| (Martin-Soelch et al., 2021) | MNI | p <.005 | History of depression>Controls: Outcome, Reward>No reward | Unspecified. |
| (Pizzagalli et al., 2009) | TAL | Uncorrected at p <.005 | Controls>MDD & MDD>Controls: Anticipation (Reward cue -No reward cue) Controls>MDD & MDD>Controls: Outcome (Gain-No change) | Unmedicated. |
| (Dichter, Kozink, McClernon, & Smoski, 2012) | MNI | Uncorrected at p <.005 | rMDD>Controls: Anticipation rMDD<Controls and Controls<rMDD: Outcome | 2 patients were receiving psychotherapy. |
| (Hall, Milne, & Macqueen, 2014) | TAL | p <.05 voxel-wise corrected | MDD>Controls & Controls>MDD: Outcome (Reward acquisition - Punishment reversal) Controls>MDD: Outcome (Large win – Small win) | 15 patients were taking antidepressants and 5 patients were taking antipsychotics. |
| (Arrondo et al., 2015) | MNI | Uncorrected | Controls>Depressed patients: Anticipation | 13 patients were taking antidepressants. |
| (Chase et al., 2013) | MNI | Cluster-wise uncorrected at p <.005 | Controls>MDD & MDD>Controls: Anticipation | 5 patients were taking antipsychotics,29 were taking |

| | | | | |
|---|---|---|---|---|
| | | | | antidepressants, 6 were taking bupropion,4 were taking mood stabilizers and 11 were taking anxiolytic. |
| (Burrows et al., 2021) | MNI | p <.05 | MDD<HC: Anticipation | 30 MDD high and 32 MDD low were taking medication. |
| (Smoski et al., 2009) | MNI | Cluster-wise corrected | MDD>HC & HC>MDD: Anticipation and Outcome | Unmedicated. |
| (Smoski, Rittenberg, & Dichter, 2011) | MNI | Uncorrected at p <.005 | MDD>HC & HC>MDD: Anticipation (Money>Images)<br>HC>MDD: Anticipation (Money)<br>MDD>HC: Outcome | 4 participants were taking antidepressants. |
| (Segarra et al., 2016) | MNI | Corrected at p <.05 | HC>MDD: Outcome | 13 depressed patients were taking antidepressant medication. |
| (Admon et al., 2015) | MNI | Uncorrected at p <.005 | HC>MDD: Outcome | Undefined. |
| (DelDonno et al., 2019) | MNI | p <.005 | HC>MDD: Anticipation | MDD participants free from any medication in the last 3 months. |
| (Gorka et al., 2014) | MNI | p <.001 | MDD>HC: Anticipation | 1 patient taking psychiatric medication. |
| (Knutson, Bhanji, Cooney, Atlas, & Gotlib, 2008) | TAL | Uncorrected at p <.016 | HC>MDD & MDD>HC: Anticipation<br>HC>MDD: Outcome | Unmedicated. |
| (Oh, Lee, Patriquin, Oldham, & Salas, 2021) | TAL | Uncorrected at p <.001 | MDD<HC: Outcome | Unmedicated. |
| (Epstein et al., 2006) | TAL | p <.05 | MDD<HC: Outcome | Unmedicated. |

| | | | | |
|---|---|---|---|---|
| (Gradin et al., 2015) | MNI | Corrected at p <.05 | HC>MDD: Outcome | Unmedicated. |
| (Keedwell, Andrew, Williams, Brammer, & Phillips, 2005) | TAL | Cluster-wise corrected at p<.01 | MDD>HC: Outcome | 11 patients were taking antidepressants. |
| (McCabe, Cowen, & Harmer, 2009) | MNI | Corrected at p <.05 | MDD<HC: Outcome | 6 patients had taken medication previously. |
| (Kumari et al., 2003) | TAL | Cluster-wise corrected at p <.005 | MDD<HC, Outcome: Positive pictures>Reference picture<br>MDD>HC, Positive pictures>Reference picture<br>MDD<HC, Positive pictures>Negative picture<br>MDD>HC, Positive pictures>Negative picture | Antidepressants and ECT previously adopted. |
| (Canli et al., 2004) | TAL | Uncorrected at p <.001 | HC>MDD, Outcome: (Happy-Neutral) | 7 patients were taking antidepressants. |
| (Fournier et al., 2013) | MNI | Uncorrected at p<.001, k>20 | MDD>HC: Outcome | 7 patients were taking antidepressants with augmentation |
| (Gotlib et al., 2005) | TAL | Uncorrected at p<.001, k>5 | MDD>Controls, Outcome: Happy-Neutral<br>Controls>MDD, Happy-Neutral | 9 patients were taking anti-depressants. |

MNI Montreal Neurological Institute, TAL Talairach, MDD Major depression disorder, HC Healthy Controls, ECT Electroconvulsive therapy

**Table S2**: PRISMA checklist 2020

| Section and topic | Item # | Checklist item | Reported on page # |
|---|---|---|---|
| **TITLE** | | | |
| Title | 1 | Identify the report as a systematic review. | 1 |
| **ABSTRACT** | | | |
| Abstract | 2 | See the PRISMA 2020 for Abstracts checklist | 2 |
| **INTRODUCTION** | | | |
| Rationale | 3 | Describe the rationale for the review in the context of existing knowledge. | 4,5 |
| Objectives | 4 | Provide an explicit statement of the objective(s) or question(s) the review addresses. | 6,8 |
| **METHODS** | | | |
| Eligibility criteria | 5 | Specify the inclusion and exclusion criteria for the review and how meta-analyses were grouped for the synthesis. | 7,8 |
| Information sources | 6 | Specify all databases, registers, websites, organisations, reference lists and other sources searched or consulted to identify studies. Specify the date when each source was last searched or consulted. | 7,8 |
| Search strategy | 7 | Present the full search strategies for all databases, registers and websites, including any filters and limits used. | 7,8 |
| Selection process | 8 | Specify the methods used to decide whether a meta-analysis met the inclusion criteria of the review, including how many reviewers screened each record and each report retrieved, whether they worked independently, and if applicable, details of automation tools used in the process. | 7,8 |
| Data collection process | 9 | Specify the methods used to collect data from reports, including how many reviewers collected data from each report, whether they worked independently, any processes for obtaining or confirming data from study investigators, and if applicable, details of automation tools used in the process | 7,8 |
| Data items | 10a | List and define all outcomes for which data were sought. Specify whether all results that | - |

| | | | |
|---|---|---|---|
| | | were compatible with each outcome domain in each study were sought (e.g. for all measures, time points, analyses), and if not, the methods used to decide which results to collect. | |
| | 10b | List and define all other variables for which data were sought (e.g. participant and intervention characteristics, funding sources). Describe any assumptions made about any missing or unclear information. | Supplementary Table 1 |
| Meta-analysis quality assessment | 11 | Specify the methods used to assess quality in the included meta-analyses, including details of the tool(s) used, how many reviewers assessed each meta-analysis and whether they worked independently, and if applicable, details of automation tools used in the process | 8,9 |
| Effect measures | 12 | Specify for each outcome the effect measure(s) (e.g. risk ratio, mean difference) used in the synthesis or presentation of results. | 10,11 |
| Synthesis methods | 13a | Describe the processes used to decide which meta-analyses were eligible for each synthesis (e.g. tabulating the study intervention characteristics and comparing against the planned groups for each synthesis. | 6 to 7 |
| | 13b | Describe any methods required to prepare the data for presentation or synthesis, such as handling of missing summary statistics, or data conversions. | Supplementary method |
| | 13c | Describe any methods used to tabulate or visually display results of individual meta-analyses. | Supplementary method |
| | 13d | Describe any methods used to synthesise results and provide a rationale for the choice(s). | 9 |
| | 13e | Describe any methods used to explore possible causes of heterogeneity among study results (e.g. subgroup analysis, meta-regression). | Supplementary method |
| | 13f | Describe any sensitivity analyses conducted to assess robustness of the synthesised results | 10 |
| Reporting bias assessment | 14 | Describe any methods used to assess risk of bias due to missing results in a synthesis (arising from reporting biases). | 10 |

| | | | |
|---|---|---|---|
| Certainty assessment | 15 | Describe any methods used to assess certainty (or confidence) in the body of evidence for an outcome. | - |
| **RESULTS** | | | |
| Study selection | 16a | Describe the results of the search and selection process, from the number of records identified in the search to the number of studies included in the review, ideally using a flow diagram. | Figure 1 |
| | 16b | Cite studies that might appear to meet the inclusion criteria, but which were excluded, and explain why they were excluded. | Figure 1 |
| Study characteristics | 17 | Cite each included study and present its characteristics. | Table 1, Supplementary table 1 |
| Quality of meta-analyses | 18 | Present assessments of quality for each included meta-analysis. | 10 to 12 |
| Results of individual meta-analyses | 19 | For all outcomes, present, for each meta-analysis: (a) summary statistics for each group (where appropriate) and (b) an effect estimate and its precision (e.g. confidence/credible interval), ideally using structured tables or plots. | 10 to 12 |
| Results of syntheses | 20a | For each synthesis, briefly summarise the characteristics and quality among contributing meta-analyses. | 10 to 12 |
| | 20b | Present results of all statistical syntheses conducted. If meta-analysis was done, present for each the summary estimate and its precision (e.g. confidence/credible interval) and measures of statistical heterogeneity. If comparing groups, describe the direction of the effect. | 10 to 12 |
| | 20c | Present results of all investigations of possible causes of heterogeneity among study results. | Table 2 and pages 10 to 12 |
| | 20d | Present results of all sensitivity analyses conducted to assess the robustness of the synthesised results. | Table2 and page14 |
| Reporting biases | 21 | Present assessments of risk of bias due to missing results (arising from reporting biases) for each synthesis assessed. | Table2 and page 14 |
| Certainty of evidence | 22 | Present assessments of certainty (or confidence) in the body of evidence for each outcome assessed. | - |

| DISCUSSION | | | |
|---|---|---|---|
| Discussion | 23a | Provide a general interpretation of the results in the context of other evidence | 14 to 20 |
| | 23b | Discuss any limitations of the evidence included in the review. | 14 to 20 |
| | 23c | Discuss any limitations of the review processes used. | 14 to 20 |
| | 23d | Discuss implications of the results for practice, policy, and future research | 14 to 20 |
| **OTHER INFORMATION** | | | |
| Registration and protocol | 24a | Provide registration information for the review, including register name and registration number, or state that the review was not registered. | 7 |
| | 24b | Indicate where the review protocol can be accessed, or state that a protocol was not prepared. | 7 |
| | 24c | Describe and explain any amendments to information provided at registration or in the protocol. | - |
| Support | 25 | Describe sources of financial or non-financial support for the review, and the role of the funders or sponsors in the review. | 20 |
| Competing interests | 26 | Declare any competing interests of review authors. | 20 |
| Availability of data, code, and other materials | 27 | Report which of the following are publicly available and where they can be found: template data collection forms; data extracted from included studies; data used for all analyses; analytic code; any other materials used in the review. | 21 |

*From*: Page MJ et al., The PRISMA 2020 statement: an updated guideline for reporting systematic reviews. BMJ 2021;372. doi: 10.1136/bmj.n71. PRISMA 2020 has been originally designed for systematic reviews of individual studies.

**Table S3.** Brief summary of the top 10 genes that were most expressed in the caudate and putamen, derived from the Brain Annotation Toolbox (BAT). The corresponding characterizations were acquired from PubMed gene.

| GENE | SUMMARY |
|---|---|
| SAG (S-antigen visual arrestin) | It is Agonist-mediated desensitization of G-protein-coupled receptors and cause specific dampening of cellular responses to stimuli such as hormones, neurotransmitters, or sensory signals. |
| MME (Membrane metalloendopeptidase) | Neutral endopeptidase that cleaves peptides at the amino side of hydrophobic residues and inactivates several peptide hormones including glucagon, enkephalins, substance P, neurotensin, oxytocin, and bradykinin. |
| SLC5A7 (Solute carrier family 5 member 7) | It encrypts sodium and chloride ion-dependent transporters. Dysregulation of this gene results in implications of disorders such as depression, attention-deficit disorder, and schizophrenia. |
| SLC35D3 (Solute carrier family 35-member D3) | It transports carbohydrate and pyrimidine nucleotide-sugar. |
| NTRK1 (neurotrophic receptor tyrosine kinase 1) | Kinase in this gene spearheads cell differentiation and it also specifies sensory neuron subtypes. |
| SFTA3 (Surfactant associated 3) | Involved in wound healing. |
| GPR101 (G protein-coupled receptor 101) | The protein encoded by this gene is an orphan G protein-coupled receptor of unknown function. |
| ZBED2 (zinc finger BED-type containing 2) | Predicted to be in the nucleus and is likely embedded in chromatin. |
| PRKAG3 (Protein kinase AMP-activated non-catalytic subunit gamma 3) | AMPK is a crucial enzyme that observes the energy status of cells as well as regulating the body’s metabolism. |
| ANKRD34B (ankyrin repeat domain 34B) | Predictions show that the gene is found with cells (cytoplasm and nucleus). |
| KCNJ1 (potassium inwardly rectifying channel subfamily J member 1) | It is triggered by internal ATP. It is thought to play a crucial role in balancing potassium levels in the body. |
| KPRP (keratinocyte proline rich protein) | This gene facilitates keratinocyte differentiation. |

A

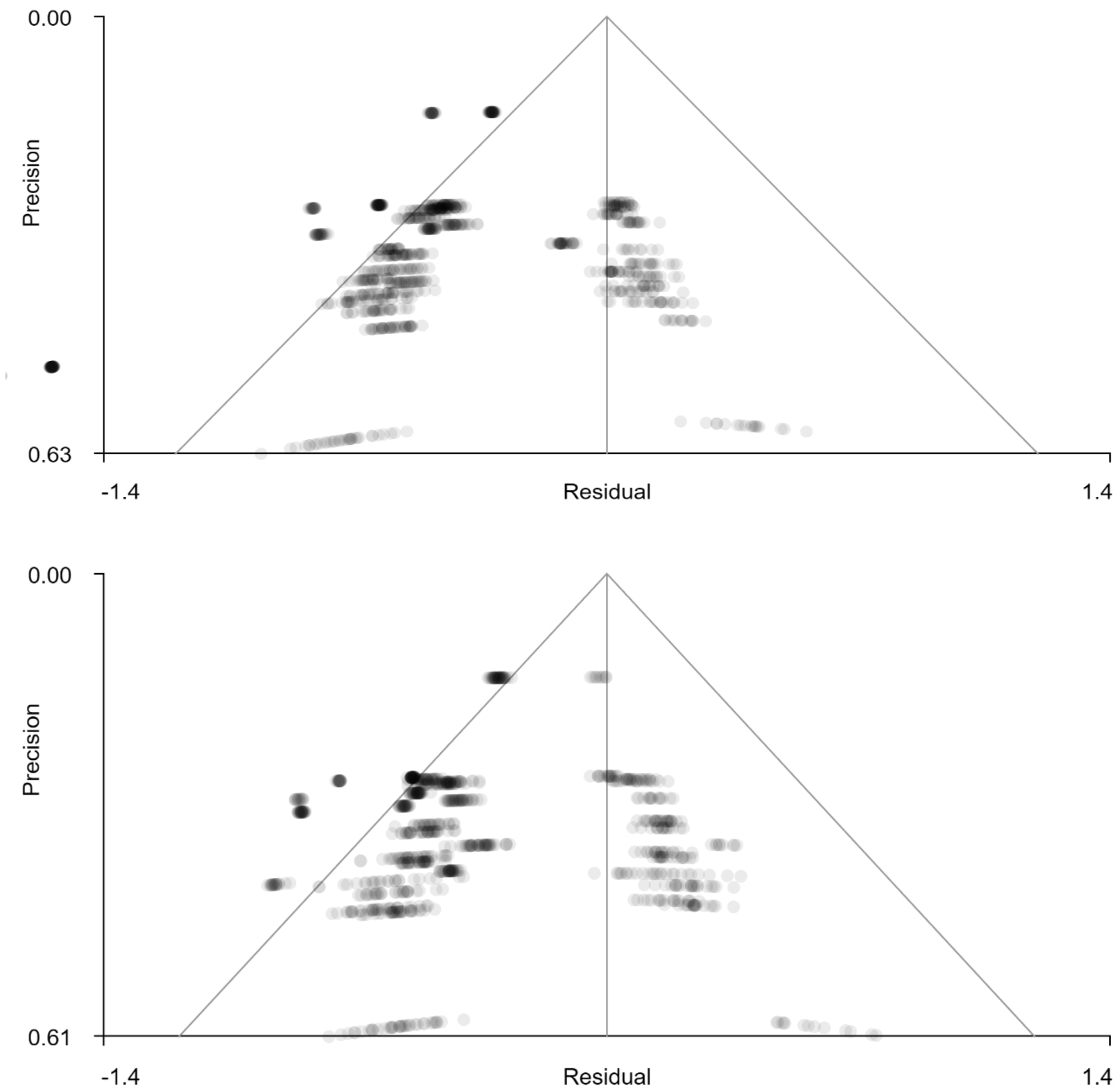
0.00
Precision
0.63
-1.4
Residual
1.4
0.00
Precision
0.61
-1.4
Residual
1.4

B

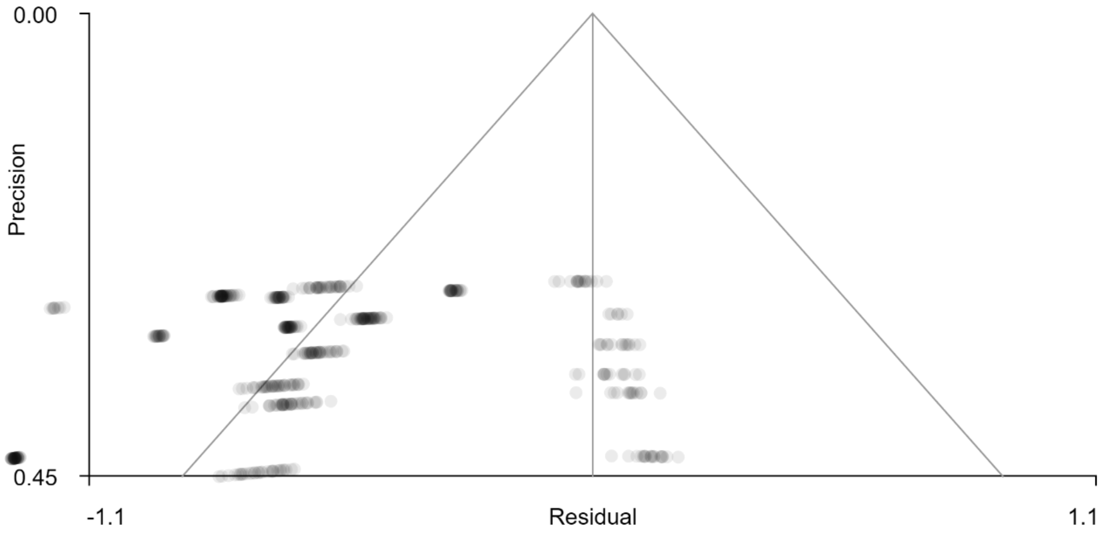
0.00
Precision
0.45
-1.1
Residual
1.1

**C**

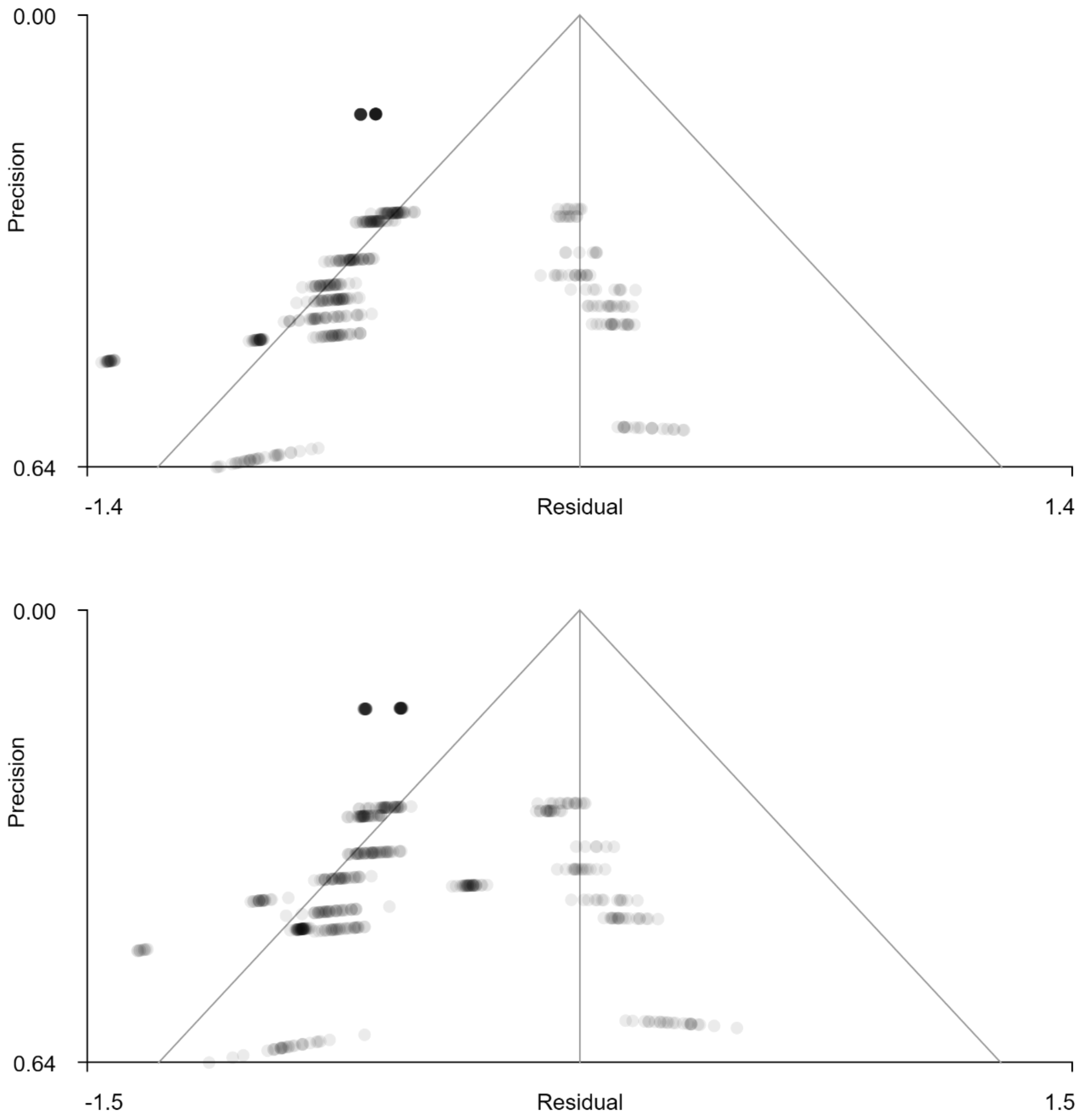
0.00
Precision
0.64
-1.4
Residual
1.4
0.00
Precision
0.64
-1.5
Residual
1.5

D

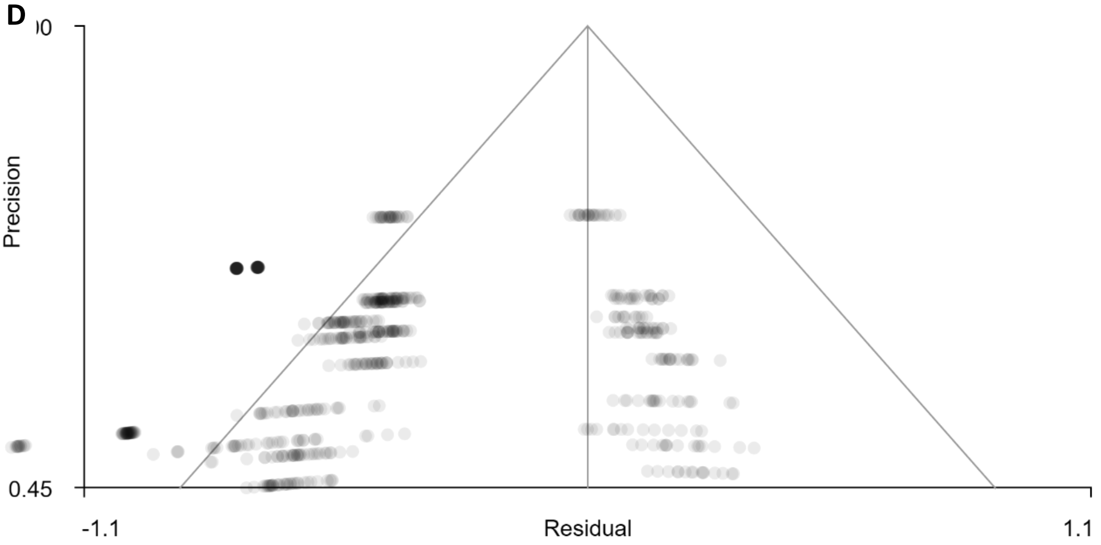


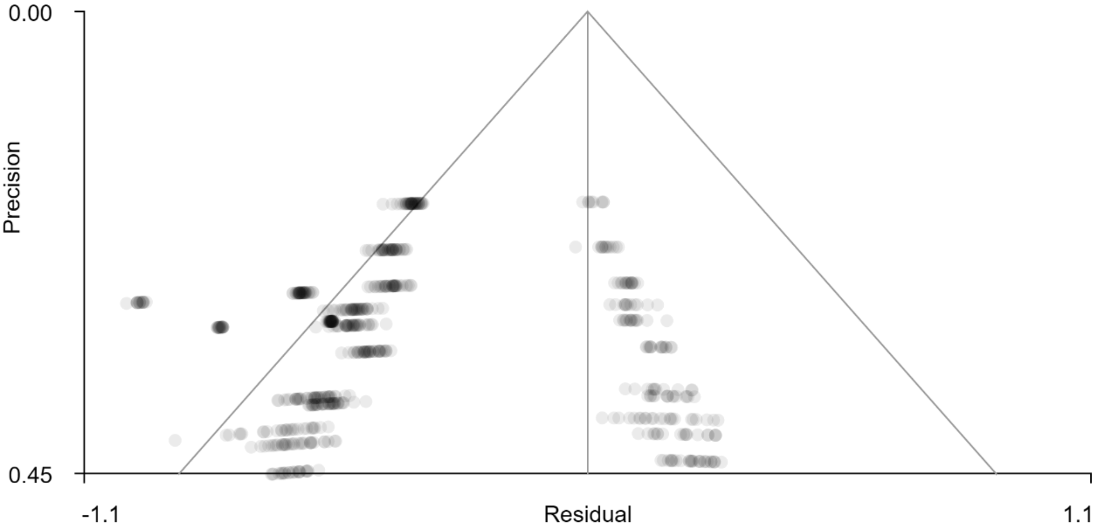


**Figure S1.** Funnel plots generated by SDM-PSI. **A** Plots for the regions identified as being altered in depression during general reward processing (top, right striatum and bottom, subgenual anterior cingulate cortex). **B** Funnel plot for monetary outcome alterations (right caudate). **C** Funnel plots for natural outcome alterations (top, right putamen and bottom, left putamen). **D** Funnel plots for monetary anticipation (top, right thalamus and bottom, left putamen).

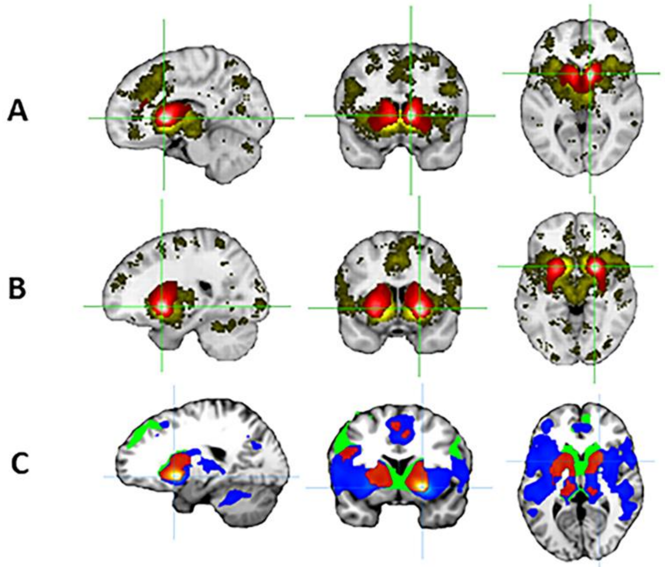


**Figure S2.** Overlap of the meta-analytic coactivation and network level patterns. **A** Functional connectivity ∩ meta-analytic co-activation of the caudate, **B** Functional connectivity ∩ meta-analytic co-activation of the putamen. **C** Voxel-based functional connectivity.

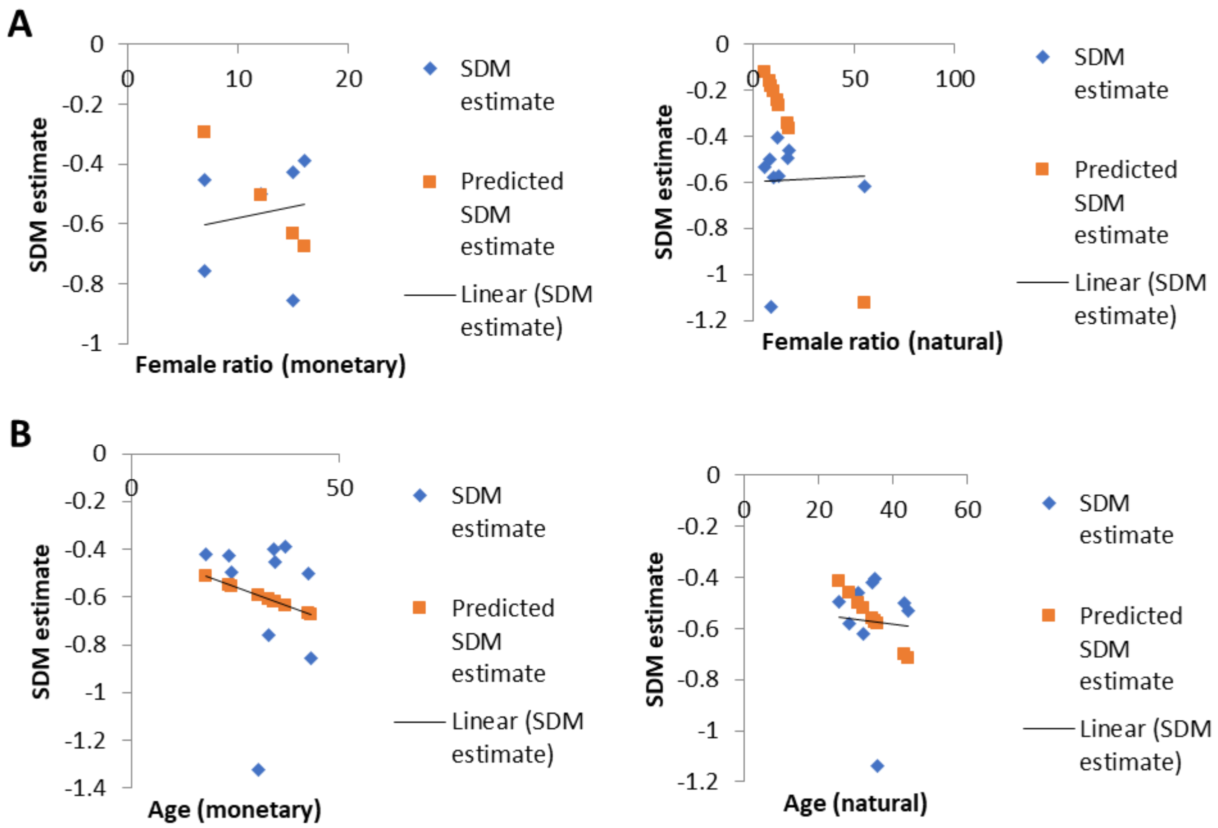


**Figure S3. A** Meta-regression plots for gender (female ratio in the patient sample, depicted on the x-axis) during monetary and natural reward outcome. **B** Meta-regression plots for age according to the two reward types.

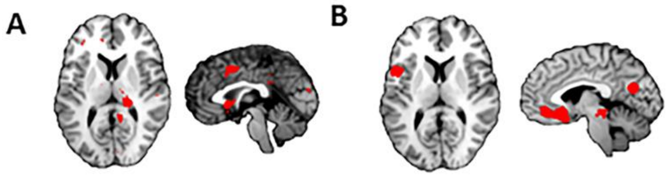


**Figure S4.** Linear model analysis of medication effect. **A** Effect on monetary reward outcome. **B** Effect on natural reward outcome.
